# The Interplay Between Breastfeeding and Autism Spectrum

**DOI:** 10.1101/2025.01.21.25320785

**Authors:** Ayelet Ben-Sasson, Aviva Mimouni-Bloch, Sukaina Samhat-Darawshi, Keren Ilann, Lidia V. Gabis

## Abstract

**Background:** Evidence supporting the association between breastfeeding patterns and ASD is inconsistent. This study examined sociodemographic and birth factors related to breastfeeding duration and ASD, compared to a typically developing (TD) cohort, using a data-driven approach based on electronic health and developmental records (EHR).

**Methods:** Demographics, feeding preferences (breastfeeding, bottle or both), breastfeeding duration as reported by parents during routine baby wellness visits, were analyzed for a cohort of 11,766 (1.9%) children with ASD and a random subsample of 12,000 (2.03%) TD children. The designation of ASD versus TD was based on a national ASD registry and assigned after EHR were completed. Pre-term, very low birth weight, multiple births, and infants with complex medical comorbidities were excluded.

**Results:** Infants with ASD were breastfed for an average of 5.0 months, 1.5 months shorter than TD. Fewer ASD infants were exclusively breastfed in the first year of life. Two-way ANOVAs indicated a significant effect of socioeconomic status (SES) and ASD on breastfeeding duration, and a significant interaction with ASD. Post-hoc comparisons showed that the Low SES ASD and TD groups were breastfed longer than all other groups (p<.001). For the Low-Medium, Medium-High and High SES groups, infants with ASD were breastfed less than TD.

**Conclusions:** Shorter breastfeeding duration among ASD was confirmed in this representative cohort, calling for closer monitoring for ASD in infants with breastfeeding difficulties. These challenges were independent of birth parameters; however, influenced by socioeconomic factors.

## Introduction

The relation between breastfeeding and autism spectrum disorder (ASD) has been explored, since breastfeeding provides a unique perspective into early social-communication cues, sensory-motor capacities, and regulatory patterns. As a shared mother-infant activity, it is inherently demanding, relying on reflexive responses and mutual coordination. Several studies have examined feeding and breastfeeding patterns, revealing that children with ASD are less likely to be breastfed, exclusively breastfed, or breastfed for an extended duration compared to typically developing (TD) children (Al-Farsi et al., 2012; da Silva et al., 2024; Elbedour et al., 2024; Schultz et al., 2006; Shafai et al., 2014; Soke et al., 2019; Tseng et al., 2019). Exclusively breastfeeding rates are significantly lower in children with ASD (12.8%) compared to TD children (36.3%) (Elbedour et al., 2024) and breastfeeding duration is often shorter by an average of two months in ASD populations (e.g., 6–8 months in ASD vs. 8.7–10 months in TD) (Elbedour et al., 2024; Peries et al., 2023; Soke et al., 2019).

Two previous studies examined whether data from national baby wellness electronic health records (EHR) could predict an ASD diagnosis. Both studies found that any duration of breastfeeding was among the top ten factors contributing to ASD prediction (Ben-Sasson et al., 2024; Ben-Sasson et al., 2024). This unexpected contribution of breastfeeding to the ASD prediction model calls for further investigation of this data.

However, this field of research is marked by contradictory results and sociodemographic inconsistencies. Some studies found no significant association between breastfeeding initiation or duration and ASD, even after controlling for confounding variables such as medical co-morbidities, birth-related factors, and sociodemographics (Abd Elmaksoud et al., 2022; Husk & Keim, 2015; Punatar et al., 2024; Say et al., 2015; Zhan et al., 2023). The conflicting results may stem from methodological differences, such as reliance on parent recall after an ASD diagnosis, inclusion of preterm infants, and varying definitions of breastfeeding measures (e.g., any vs. exclusive breastfeeding). Additionally, factors known to independently challenge breastfeeding, such as preterm birth or maternal health, are not accounted for consistently. Studies using large, epidemiologically representative cohorts that control for confounders and isolate factors affecting breastfeeding are needed to address these discrepancies. These efforts could clarify the nature of the relationship between breastfeeding duration and ASD, potentially uncovering early behavioral markers and possible venues for early intervention.

Infant feeding practices are influenced by healthcare, economic, social and cultural contexts. In the Israeli population, 53.9% of women are exclusively breastfeeding at two months postpartum. This drops to 15.3% by six months (Israel Ministry of Health, 2023). Globally, the rate of exclusive breastfeeding up to age six months was about 25% in 2024 (Centers for Disease Control and Prevention, 2024). Breastfeeding duration tends to be shorter in high-income countries compared to those that are resource-poor (Victora et al., 2016). At the same time, the probability of exclusive breastfeeding is lower for mothers who have less education, are less religious, and are younger (Gabay, et al., 2022, Reynolds et al., 2023). These findings highlight the mixed evidence regarding sociodemographic factors and breastfeeding likelihood.

Breastfeeding difficulties in infants with ASD can arise from various infant and maternal factors. For infants, these may include challenging feeding behaviors such as dysregulated vigorous sucking due to sensory regulation issues or weak sucking due to motor difficulties that may lead to prolonged feeding duration, weaning difficulties and refusal (Allen et al., 2019; Ghozy et al., 2020; Lucas & Cutler, 2015; Varma & de Souza, 2023). Feeding challenges, including food selectivity and inadequate intake, have been observed in infants with subsequent diagnoses of ASD, which may contribute to breastfeeding refusal (van ’t Hof et al., 2020; Soke et al., 2019). Moreover, genetic syndromes related to ASD, such as those presenting with insulin-like growth factor-1 deficiency, failure to thrive and orofacial dysfunction, can further exacerbate breastfeeding difficulties. Reduced eye contact during feeding and difficulty communicating hunger cues may be linked to early signs of ASD (Kara & Alpgan, 2020).

Some mothers of ASD children may have a higher likelihood for ASD traits or autism diagnosis. This group may face unique breastfeeding obstacles, including sensory sensitivities, communication differences, clumsiness, and difficulties with executive function (Grant et al., 2022; Wilson & Andrassy, 2022). These can complicate breastfeeding despite extensive knowledge and determination to breastfeed (Eidelman, 2022).

Evidence is inconsistent whether demographic factors contribute to breastfeeding patterns in ASD. Peries et al. (2023) reported higher rates of breastfeeding among mothers with higher levels of education and socioeconomic status (SES). In a recent study from Israel, that included Jewish and Bedouin children, ethnicity did not impact breastfeeding patterns of children with ASD during the first year of life (Elbedour et al., 2024). This was also shown in ASD studies, once demographics (e.g., ethnicity, parent education, mother’s age and income) were controlled for (da Silva et al., 2024; Husk & Keim, 2015; Zhan et al., 2023) and for breastfeeding initiation (Soke et al., 2019). Nonetheless, some studies that link ASD with breastfeeding likelihood do not consider demographic moderators such as SES (Aloufi, et al., 2022; Huang, et al., 2021; Ghozy, et al., 2020; Shafai, et al., 2014).

Additional factors shown to reduce the likelihood and duration of breastfeeding were detrimental birth parameters, including Cesarean delivery, multiple births, prematurity, and low birth weight (Benetou et al., 2019; Shani & Shinwell, 2003; Spyrakou et al., 2022; Zimmerman et al., 2022). Breastfeeding a premature infant presents unique challenges and success rates are generally low, especially in neonatal units (Nascimento & Issler, 2004). Given that children with ASD exhibit higher rates of premature birth (Limperopoulos, 2009; Ouss-Ryngaert et al., 2012), Cesarean section and multiple births (Kolevzon et al., 2007), these factors likely contribute to the breastfeeding difficulties and may be confounding factors. However, birth parameters are not addressed consistently in research on breastfeeding and ASD (Schultz et al., 2006; Xiang et al., 2023). When birth parameters like prematurity were controlled for, significant differences in breastfeeding patterns between ASD and TD groups remained, indicating that birth-related factors alone do not fully account for these disparities (Tseng et al., 2019; Varma & de Souza, 2023). The current study examined the relation between ASD and breastfeeding patterns, considering sociodemographic and birth parameters in a large-scale database.

## Methods

### Study Design and Ethics

This population-based, EHR study was approved by the Helsinki Committee of the Israeli Ministry of Health (MoH) (#15/2021). Data was extracted and anonymized using TIMNA, which was established by the MoH for conducting big data health research. Data anonymization and security were managed by the TIMNA team, ensuring the study data did not contain any identifying information.

### Data Source and Measurement

The EHRs analyzed in this study reflected a cohort followed by MoH well-baby clinics. The MoH is the main provider of developmental check-ups at 503 clinics across Israel, covering 75% of the population. The services are provided by nurses with specific training in maternal and child health and by a developmental pediatrician. In each of the 11 visits from birth to 6 years, providers deliver immunizations, survey growth and development, and identify medical and environmental risks to the mother and child. The Edinburg postpartum depression survey (Cox et al., 1987) is also administered.

Additionally, the clinics aim to promote a healthy lifestyle and prevent diseases through guidance on nutrition, breastfeeding, child development, hygiene, and safety (Israeli Ministry of Health, 2024). At each visit, nurses query the parent about the child’s feeding: Is the child breastfed? Fed by formula? When did breastfeeding end? (Israeli Ministry of Health, 2017). For this study, the child’s feeding data was analyzed in detail.

### Participants

Inclusion criteria for EHR extraction were children born from January 2014 through December 2019, with at least 2 years of baby wellness surveillance in the Israeli MoH database. See Figure 1 for flow diagram of participants.

**Figure 1.**
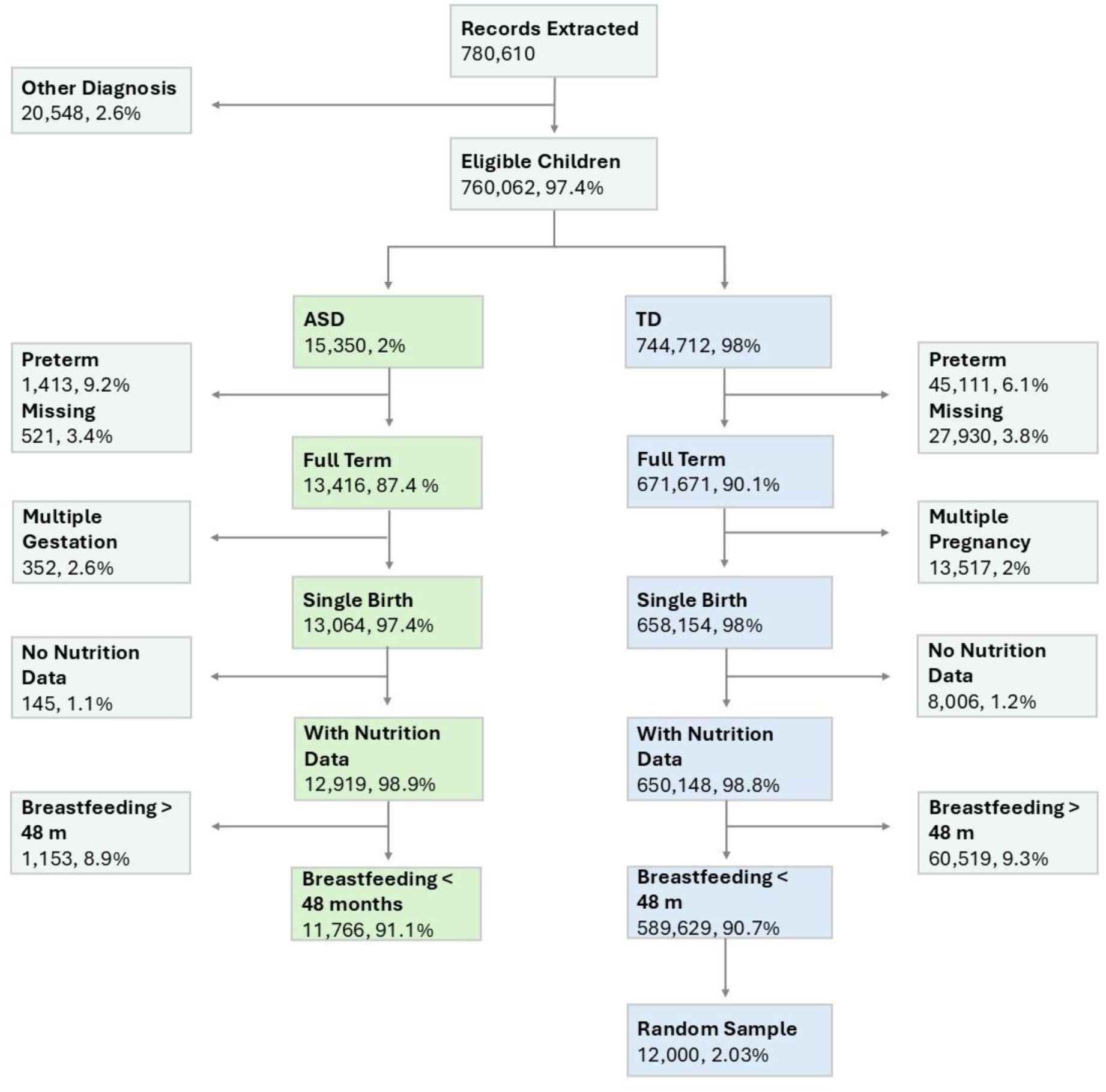
Flow Diagram

ASD diagnosis status was based on receiving benefits from the Israel National Social Security for an ASD diagnosis. Diagnosis requires psychiatric/neurodevelopmental, neurological or developmental pediatrician assessments, standard intelligence/developmental test, and results of a formal ASD diagnostic tool. This data was extracted from the Social Security database and cross-referenced with the MoH database by TIMNA.

Infants without breastfeeding data and those with breastfeeding duration over 48 months were also excluded, as were excluded infants with health conditions that may impair or prevent breastfeeding, such as cleft palate, chromosomal anomalies, choanal atresia, etc. To minimize the role of known birth factors linked to ASD and breastfeeding, prematurity, birth weight less than 1,750 grams and infants from multiple gestations were excluded as well. Finally, a randomly selected sample of 12,000 TD infants was selected.

### Data Analysis

The data was analyzed using SPSS software, version 27 (IBM Corp., Armonk, NY, USA). Clinical nutrition data was engineered to enable its processing. This included managing multiple records per child over time. The log of this data consisted of the date of the child’s visit, whether the child was breastfeeding, and whether the child was fed with formula. For each child, we computed breastfeeding duration in months, and feeding type for three age ranges^1^: 0-3, 3-6, and 6-12 months. Feeding types were classified as exclusively breastfed, mixed breastfeeding and formula or formula exclusively, and were computed based on the last nutrition datapoint within each age range. The last nutrition datapoint available per child was at an average of 8.5 months, with a wide standard deviation (SD) of 7.8.

SES was based on the classification of the geographic location of the family’s well-baby clinic into 10 clusters by the Central Bureau of Statistics. For the current study, the 10 clusters were grouped into 4 SES groups: Low SES, Low-Medium SES, Medium-High SES, High SES.

All continuous variables (breastfeeding duration, birth weight, gestational age at birth, maternal age at birth) showed skewness and kurtosis values under 2, suggesting that the distributions were approximately normal. Hence, parametric tests were conducted for further analyses. Differences between the randomly and non-randomly selected groups were non-significant using chi-square testing, to ensure non-bias on key variables (gender, ethnicity, mother’s country of origin, type of education, and SES status). In addition, the randomly selected group did not differ from the rest of the sample in breastfeeding duration.

To determine which potential confounding variables (i.e., demographics and birth-related) should be integrated in the main models, the relationships between breastfeeding duration and these variables were examined using correlations or chi-square tests.

Two-way ANOVA models compared breastfeeding duration between groups considering the background variables of ethnicity, delivery type, maternal age group, and SES group. The models were separated for each background variable because sample size varied as a factor of available data for each of these variables (see Table 1).

**Table 1.** Child and Family Demographics, Birth and Postnatal Parameters of the TD and ASD Groups.

| Characteristic | TD (n=12,000) | ASD (n=11,766) | Statistics<sup>a</sup> |
| --- | --- | --- | --- |
| Male, n (%) | 8,995 (50.5%) | 6,055 (76.4%) | p<.001, $\Phi$ =-.27, OR=.31 |
| Mother's age (years) | | | $\chi^2(2)$ =42.89 ***, V=.04 |
| ≤20 | 347 (2.9%) | 303 (2.6%) |  |
| 20.5-40 | 11,136 (92.8%) | 10,722 (91.1%) |  |
| >40 | 398 (3.3%) | 668 (5.7%) |  |
| Missing | 119 (1%) | 73 (0.6%) |  |
| Child ethnicity | | | p<.001, $\Phi$ =-.15, OR=.50 |
| Jewish | 7379 (61.5%) | 8476 (72%) |  |
| Other<sup>b</sup> | 3398 (28.3%) | 1967 (16.7%) |  |
| Missing | 212 (10.2%) | 182 (11.2%) |  |
| Mother birth country | | | p<.001, $\Phi$ =-.03, OR=.79 |
| Israel | 9922 (82.7%) | 8637 (73.4%) |  |
| Other | 1259 (10.5%) | 2294 (19.5%) |  |
| Missing | 819 (6.8%) | 835 (7.1%) |  |
| Mother's employment status | | | $\chi^2(2)$ =77.38 ***, V=.07 |
| Employed | 5149 (43.3%) | 5488 (46.6%) |  |
| Unemployed | 2525 (21%) | 2243 (19.1%) |  |
| Student | 510 (4.3%) | 299 (2.5%) |  |
| Missing | 3771 (31.4%) | 3736 (31.8%) |  |
| SES status | | | $\chi^2(3)$ =361.96 ***, V=.14 |
| Low | 3092 (25.8%) | 2222 (18.9%) |  |
| Low-medium | 2226 (18.6%) | 2995 (25.5%) |  |
| Medium-high | 1874 (15.6%) | 2661 (22.6%) |  |
| High | 1193 (9.9%) | 1281 (10.9%) |  |
| Missing | 3615 (30.1%) | 2607 (22.2%) |  |
| Birth weight, kg; mean (SD, min-max) | 3.306 (0.43, 1.835-5.314) | 3.312 (0.446, 1.81-5.43) | t (23764) = -.94, p = .347, d = .014 |
| Pregnancy week, mean (SD, min-max) | 39.494 (1.64, 37-43) | 39.346 (1.2, 37-43) | t (23764) = 9.60***, d = .102 |
| Birth type | | | $\chi^2(2) = 189.68$ ***, V = .09 |
| Spontaneous | 8958 (74.7%) | 7940 (67.5%) |  |
| Cesarean | 1767 (14.7%) | 2469 (21%) |  |
| Instrumental | 641 (5.3%) | 779 (6.6%) |  |
| Missing | 643 (5.3%) | 578 (4.9%) |  |

*Note.* Missing values were not included in the above analyses.
<sup>a</sup>Categorical variables were analyzed using Chi-square tests, binary variables using Fisher's exact tests and continuous variables using independent samples t-test. <sup>b</sup>Arab Muslim, Druze, Muslim Bedouin and Arab Christian.
\*\*\* p < .001

Chi-square tests within each SES group compared the ASD and TD groups for their rates of feeding types within the three age periods (12 tests). Adjusted z-tests tested pairwise differences between groups per feeding type within age group and SES class. Bonferroni correction for multiple comparisons was applied so that the p-value threshold was set to .001.

## Results

### Descriptives

Figure 1 shows the flow of the sample inclusion process. Initially, the sample included 780,610 children. After applying exclusion criteria to both cohorts, the final sample comprised 589,629 (98.1%) TD controls and 11,766 (1.9%) with ASD diagnosis. To account for cohort size, a random subsample of 12,000 (2.03%) TD children was selected.

Table 1 presents demographic, birth and postnatal differences between the ASD and TD groups. There were significantly more males, older mothers, Jewish families, immigrant mothers, employed mothers, and those of higher SES in the ASD group. Although children born prematurely were excluded, the ASD group had a significantly shorter pregnancy duration and higher rates of Cesarean and instrumental deliveries. About 25% of mothers met the clinical criteria for postpartum depression but there were no significant differences in the rates of postpartum depression between groups (χ²(3)=1.71, *p*=.635, *V*=.10).

### Comparison of breastfeeding duration between groups

The ASD group had a significantly (*t* (23764) =19.13, *p*<.001, *d*=.25) shorter duration of breastfeeding of 5.0 months (SD=6.10, 0-42 months) versus 6.6 months in the TD group (SD=6.30, 0-48 months). A closer examination was made possible by looking at the feeding methods of exclusively breastfed, exclusively formula fed, and mixed formula and breastfed, within three age ranges: 0-3, 3-6, 6-12 months (see Figure 2). The most dramatic differences between the ASD and TD groups were observed for rates of exclusive breastfeeding, with 24.98% vs. 37.85% at 0-3 months, 20.03% vs. 29.19% at 3-6 months and 12.20% vs. 19.65% for 6-12 months, respectively.

**Figure 2.**
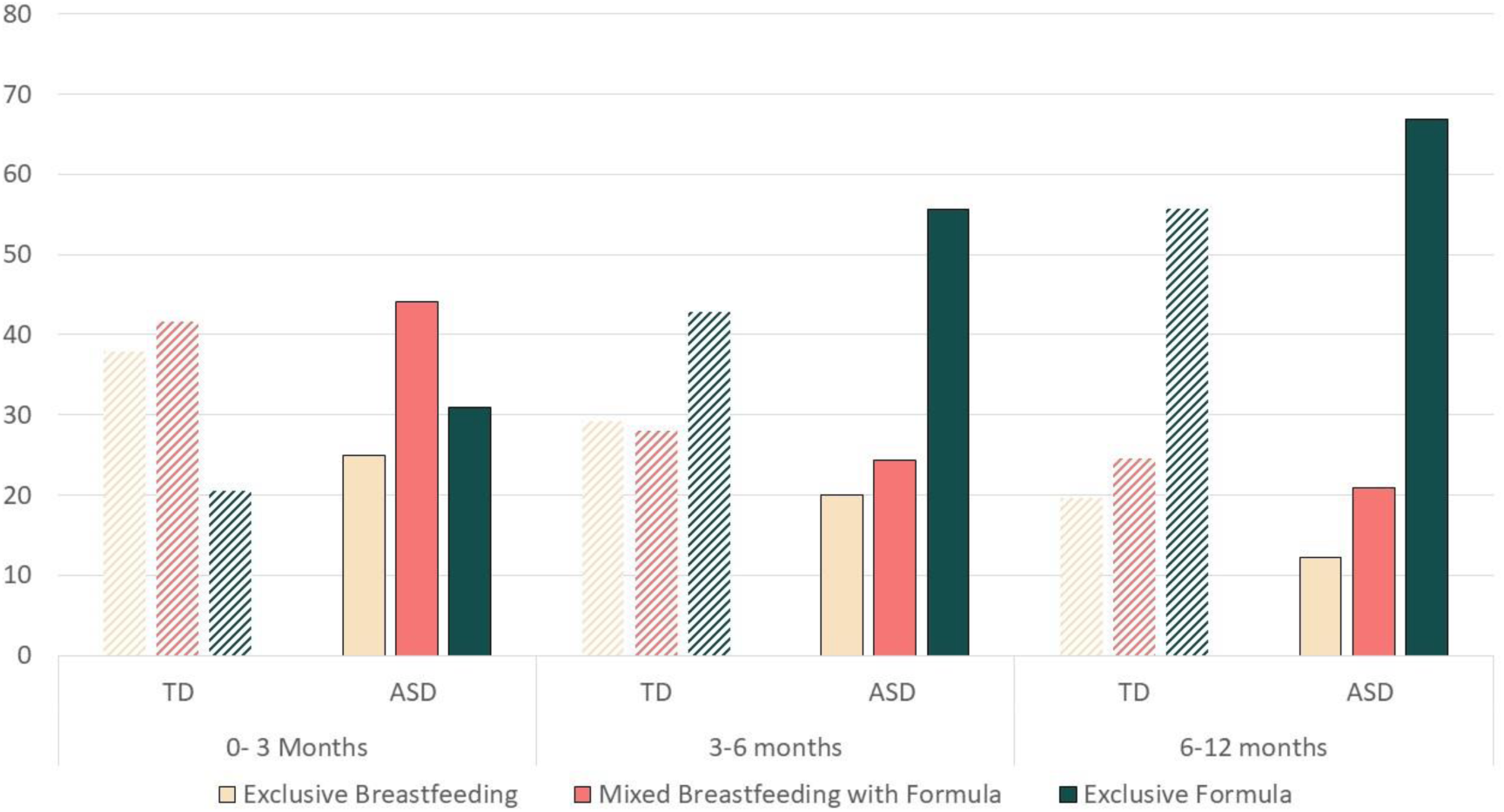
Percentage of Children in Each Feeding Category by Age

### The relation between breastfeeding duration and confounding variables

For the demographic variables in the TD population, breastfeeding duration was shorter between Jewish (5.6 months ± 6.2) and non-Jewish families (6.4 months ± 6.3), *t*(21218) = −7.40, *p* < .001, *d* = .12. Moreover, a significant difference in breastfeeding duration according to SES was found (*F*(3, 17,540) = 443.41, *p* < .001, ηp² = .07). Children in the Low SES group were breastfed for the longest period (8.3 months ± 6.1), while children in the High SES group were breastfed for an average of 6.0 months (± 6.4), Low-Medium SES (4.5 months ± 6.1) and Medium-High SES (4.3 months ± 5.8) had shorter breastfeeding durations. Looking at the three feeding type periods (0-3, 3-6, 6-12 months), there was a significant association between feeding types and SES across age groups (ꭓ2(6) = 961.77, *p*<.001, V =.17; ꭓ2(6) = 1075.27, *p*<.001, V = .19; ꭓ2(6) = 1017.87, *p*<.001, V = .18, respectively). Follow-up adjusted Z-tests indicated that the Low SES group had significantly higher rates of exclusive breastfeeding than all other groups. The High SES group had significantly higher rates of exclusive breastfeeding than both of the middle SES groups.

Regarding birth variables, a significant difference was found in breastfeeding duration by type of delivery (*F*(2, 22,551) = 169.6, *p* < .001, ηp² = .015). Children born via spontaneous delivery were breastfed for the longest period (6.3 months ± 6.3), compared to instrumental delivery (5.4 months ± 6.2), and Cesarean delivery (4.3 months ± 5.8). A significant difference in breastfeeding duration by maternal age was also observed (*F*(2, 23,571) = 8.25, *p* < .001, ηp² = .001). Mothers over 40 years of age breastfed for the shortest period (5.1 months ± 6.8), while mothers ages 20 to 40 breastfed for an average of 5.9 months (± 6.2), and those under 20 breastfed for an average of 5.8 months (± 5.5).

### Two-way ANOVA for breastfeeding duration between groups

Four, two-way ANOVAs were conducted to assess differences in breastfeeding duration between the ASD and TD groups, considering the key demographic (SES status, ethnicity) and birth variables (birth type, mother’s age group) associated with breastfeeding. Table 2 shows significant effects for the ASD group and the background variable across models. See Figure S1 for mean duration differences between groups. The interaction effect was significant only for the ASD by SES group in the ANOVA (*F*(3, 17,536)=11.46, *p*<.001, *η_p_²*=.002; see Figure 3).

**Figure 3.**
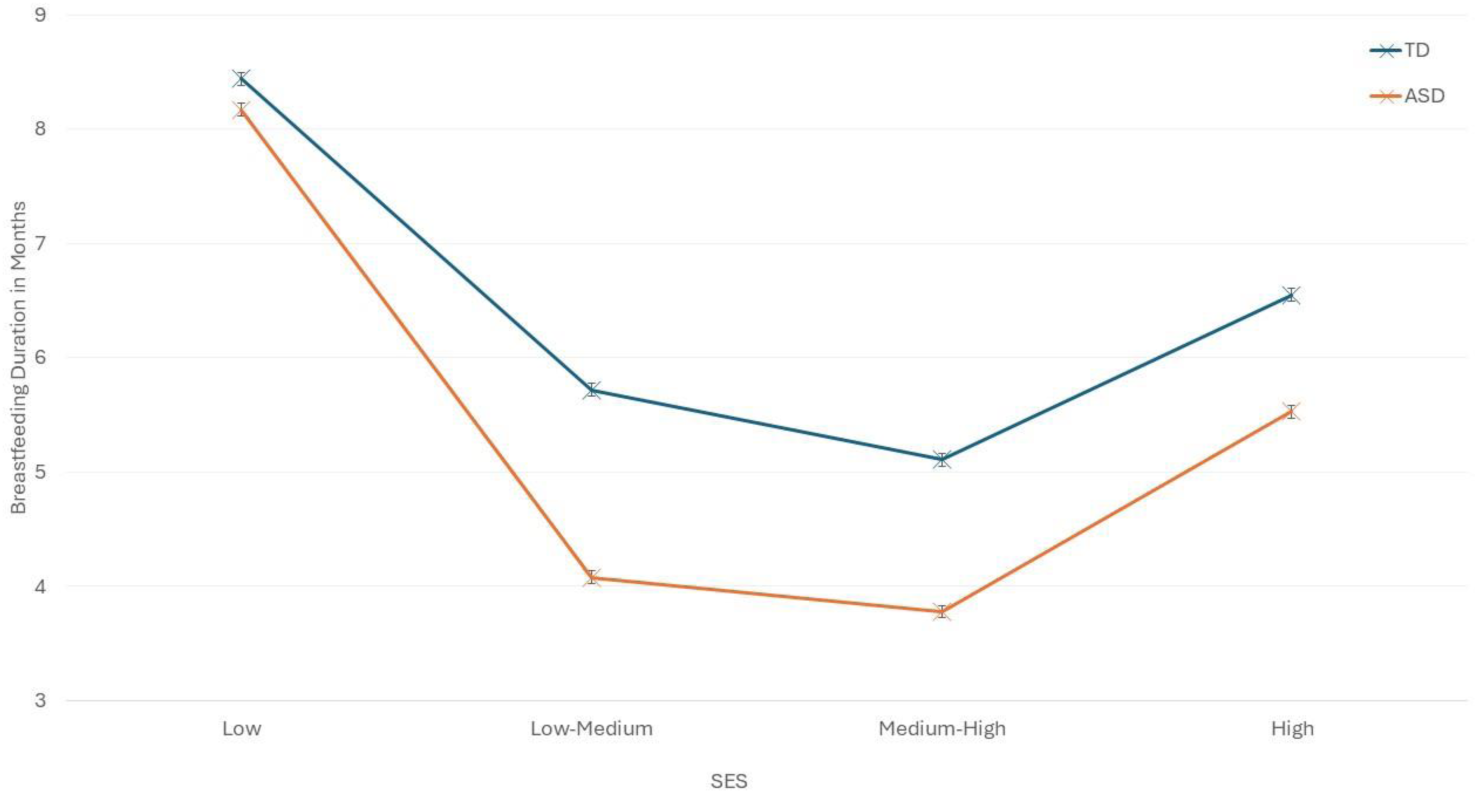
Mean and SE of Breastfeeding Duration by SES

**Table 2.** ANCOVA of Breastfeeding Duration and ASD Covarying for Demographic Variables.

| Variable | Mean (SD) |  | *F(df), $\eta_p^2$* |  |  |
| --- | --- | --- | --- | --- | --- |
|  | TD<br>(n=12,000) | ASD<br>(n=11,766) | ASD Group<br>Effect | Background<br>Variable Effect | Interaction<br>Effect |
| Child's ethnicity |  |  |  |  |  |
| Jewish | 6.49 (6.26) | 4.91 (5.98) | 241.25 (1) *** | 21.69 (1)*** | .05 (1), .000 |
| Other<sup>a</sup> | 6.93 (6.30) | 5.39 (6.19) |  |  |  |
| SES status |  |  | 124.86 (1)***,<br>.007 | 403.03 (2)***,<br>.065 | 11.46 (2)***,<br>.002 |
| Low | 8.44 (6.07) | 8.17 (6.19) |  |  |  |
| Low-Medium | 5.72 (6.22) | 4.08 (5.86) |  |  |  |
| Medium-High | 5.11 (6.07) | 3.78 (5.51) |  |  |  |
| High | 6.55 (6.31) | 5.53 (6.17) |  |  |  |
| Birth type |  |  | 126.78 (1)***,<br>.006 | 141.05 (3)***,<br>.012 | .12 (3), .000 |
| Spontaneous | 6.94 (6.28) | 5.50 (6.21) |  |  |  |
| Cesarean | 5.13 (6.19) | 4.32 (5.82) |  |  |  |
| Instrumental | 6.29 (6.25) | 5.43 (6.24) |  |  |  |
| Mother's age<br>(years) |  |  | 70.73 (1)***,<br>.003 | 403.03 (2)***,<br><.001 | 2.10 (2), .000 |
| ≤20 | 6.52 (5.84) | 4.99 (5.06) |  |  |  |
| 20.5-40 | 6.60 (6.26) | 5.12 (6.12) |  |  |  |
| >40 | 6.52 (7.57) | 4.22 (6.08) |  |  |  |

*Note.* Sample size differed for each background variable based on the missing data rates for that variable see Table 1. \*\*\* $p < .001$
<sup>a</sup>Arab Muslim, Druze, Muslim Bedouin and Arab Christian.

### Interaction effect between SES and ASD

Follow-up analyses were conducted to understand the ASD by SES interaction effect. Figure 3 shows the mean duration of breastfeeding differences between the eight SES groups (Low ASD, Low TD, Low-Medium ASD, Low-Medium TD, Medium-High TD, Medium-High ASD, High TD, and High ASD). ANOVA showed a significant group effect *F*(7, 17,536)=216.1, *p*<.001, *η_p_²*=.079. Bonferroni post-hoc tests revealed a non-significant difference between the following subgroups: Low ASD versus Low TD (both with a longer average breastfeeding durations). Other comparisons within SES groups were all significant (p < .001), with shorter breastfeeding durations for ASD versus TD.

For each of the three age groups, chi-square analyses compared rates of feeding types between groups within each SES group (see Figure S2a-2c). <u>For age 0-3 months</u>, Chi-square tests were significant for all SES groups but with a lower effect size for the Low SES group (Low: ꭓ2(2) = 37.79, *p*<.001, V = .09; Low-Medium: ꭓ2(2) = 193.4, *p*<.001, V = .20; Medium-High: ꭓ2(2) = 82.58, *p*<.001, V = .14; and High: ꭓ2(2) = 53.09, *p*<.001, V = .15). Pairwise comparisons were significant between groups for all feeding types. In <u>the 3-6 months period</u>, Chi-square tests were significant for all but the Low SES group (Low: ꭓ2(2) = 12.86, *p=*.002, V = .05; Low-Medium: ꭓ2(2) = 127.58, *p*<.001, V = .16; Medium-High: ꭓ2(2) = 53.94, *p*<.001, V = .11; High: ꭓ2(2) = 39.36, *p*<.001, V = .13). Pairwise comparisons within the three significant SES groups were significant for exclusive breastfeeding and exclusive formula. <u>In the 6-12 months period</u>, Chi-square tests were significant for all but the Low SES (Low: ꭓ2(2) = 3.24, *p*=.20, V = .02; Low-Medium: ꭓ2(2) = 121.6, *p*<.001, V = .15; Medium-High: ꭓ2(2) = 63.16, *p*<.001, V = .12, High: ꭓ2(2) = 30.19, *p*<.001, V = .11). The High SES group had no significant difference for the mixed feeding type, while the middle groups showed significantly lower rates of exclusive breastfeeding for ASD versus TD, versus higher rates of exclusive formula and mixed feeding for the ASD versus TD groups.

## Discussion

This EHR-based research allowed for the analysis of a large, representative birth cohort of children in Israel. By using documented feeding practices from birth and labeling records based on subsequent ASD diagnoses, it enabled a comparison of breastfeeding duration between children subsequently diagnosed with ASD to children with TD. The key finding was that children with ASD tend to be breastfed for shorter periods compared to their neurotypical peers; in support of studies in the field (Schults et al, 2016; Shafai et al, 2014; Soke et al, 2019; Tseng et al, 2019). Moreover, rates of exclusive breastfeeding were lower, and rates of exclusive formula use were higher in ASD compared to TD infants across age groups, during the first year of life, consistent with previous evidence (Al-Farsi et al., 2012; da Silva et al., 2024; Elbedour et al., 2024). The differences between ASD and TD infants in our study were observed even after the elimination of potentially confounding factors, including medical complications, premature births and multiple births. While a myriad of birth (e.g., delivery type, gestational age at delivery, maternal age) and demographic (e.g., ethnicity) parameters were associated with breastfeeding, only SES contributed to breastfeeding patterns in ASD.

Breastfeeding is a developmentally complex crosstalk between mother and infant, which requires integration of sensory input, complex motor function, and communication between the two parties. Breastfeeding also demands a mutual, persistent engagement that can be frustrating for both mother and infant. As such, it is perhaps expected that children with ASD, as a group, tend to be breastfed for shorter durations. In previous ASD studies, the difference in breastfeeding duration between ASD and TD groups was approximately 2 months (Elbedour et al, 2024, Soke et al. 2019, Xiang et al. 2023), whereas in the present study, the difference was 1.5 months. This smaller difference may be attributed to the exclusion of infants with health conditions (e.g., cleft palate, chromosomal anomalies) or birth factors (e.g., prematurity, multiple births, birth weight under 1750 grams) that could hinder or prevent breastfeeding regardless of ASD. This supports the understanding that medical factors contribute to some degree to the lower rates of breastfeeding in ASD.

In the current nation-wide study, the rate of any breastfeeding in the ASD group (67%) was notably higher than that observed in a recent study based in the southern region of Israel (47.2%; Elbedour et al., 2024). However, the rate of any breastfeeding in the TD group (77%) was similar in the studies (75.6%; Elbedour et al, 2024). The differences in rates of ASD breastfeeding between studies may relate to the conservative inclusion criteria in our study. Scrutinizing the crude measure of any breastfeeding shows that the rate of exclusive breastfeeding up to 3 months in the ASD group was 24.98% versus 37.85% in the TD group. The gap between the two groups decreased in the 3-6-month age group, with 20.03% of the ASD group exclusively breastfed versus 29.19% in the TD group. During ages 6-12 months, reported rates were 12.20% and 19.65% in the two groups, respectively.

This study reinforces the association between ASD and shorter breastfeeding duration. However, we did not measure the myriad of maternal traits and infant-related symptoms that could explain this relationship. Various aspects of ASD, including infant dysregulation, impaired social communication, maternal ASD traits and infant-mother synchrony, may contribute to the lower rates of breastfeeding observed (Berthoz et al., 2013; Henderson et al., 2003, Ingersoll & Hambrick, 2011). Motor delays and hypotonia that are characteristic of some children with ASD (Gabis et al., 2021) may lead to difficulties initiating and maintaining breastfeeding, which may partially explain the shorter duration of breastfeeding found in our study. Nonetheless, the exclusion of children with poor birth outcomes and medically complex cases reduced the likelihood of significant motor delays. Thus, it is plausible that the exclusion of premature infants and those with medical comorbidities led to a smaller gap in breastfeeding duration and hence, to non-significant interactions between birth parameters and ASD. It is therefore, not surprising that other contributing factors such as SES were revealed, as discussed further below. Regardless of the source of these difficulties–be it the child, the mother, or their synchrony– compromised breastfeeding patterns may undermine the protective advantages that breastfeeding offers (Krol et al, 2015; Soke et al, 2019) in infants later diagnosed with ASD.

In the interaction between SES and breastfeeding, mothers with low SES tended to breastfeed their infants for a longer duration regardless of a later ASD diagnosis of their children. This is in line with the longer breastfeeding in low-versus high-income countries (Victora et al., 2016). Moreover, closer investigation of exclusive breastfeeding by age group showed that among those with Low SES, rates were similar between groups. We suggest that in this lower income population, the economic pressure to breastfeed rather than formula feed may be a stronger motivation than facing the difficulties of a challenging infant. When mothers in the Low-Medium and Medium-High SES groups face difficulties breastfeeding, they may be more able to afford the cost of formula. We assume that some of the mothers in the Low-Medium and Medium-High SES groups return to work after several months, making it more difficult for them to maintain breastfeeding with a challenging infant. In contrast, in the Low SES group, the rate of unemployed mothers, who are more likely to continue breastfeeding might be higher. This could explain the lack of difference in breastfeeding duration found between the ASD and TD groups.

Factors commonly associated with ASD, such as male sex, advanced maternal age, and Cesarean delivery did not explain the breastfeeding differences between ASD and TD infants observed in our study. While these factors significantly differentiated the two groups and showed independent effects on breastfeeding, they did not interact with ASD. This reinforces the notion that shorter breastfeeding duration may serve as an early, independent risk marker for a later ASD diagnosis.

The present study introduced key new concepts. Unlike prior research that relied on parent-reported ASD or limited samples, we used a large, representative cohort with ASD diagnoses confirmed through multidisciplinary evaluations. Population-based health records allowed us to consider diverse sociodemographic and medical factors, and to exclude children with conditions like prematurity, which impede breastfeeding and increase ASD risk. Frequent well-baby visits ensured breastfeeding data was collected promptly, minimizing recall bias and avoiding diagnostic bias, since it was collected prospectively before ASD diagnosis; thus, enhancing the accuracy and reliability of our findings.

### Limitations

This study was limited in that it did not include specific data on the quantity or frequency of breastfeeding and formula intake. Instead, we inferred breastfeeding duration from multiple timepoints. Additionally, the mixed feeding category in our study included all infants who had at least one entry of breastfeeding and one entry of formula feeding in the system. This made this category less informative as it was impossible to distinguish between individuals who were mainly breastfed and those who were mainly formula fed. The interaction between SES and breastfeeding was driven mainly by the exclusive breastfeeding and exclusive formula feeding groups. This is not surprising, as the mixed feeding category included a wide spectrum of feeding patterns, and its results were not discussed in detail. The study also lacked specific data regarding underlying factors explaining the lack of or shorter periods of breastfeeding. Lastly, many of the EHR lacked paternal data and information on maternal education and employment status. Therefore, we could not use this data to better understand the SES ASD interaction.

### Conclusion

This study stemmed from a broader study that identified that having been ever breastfed was one of the top ten predictors of ASD in EHR, along with developmental milestones, birth factors, and physical growth (Ben-Sasson et al., 2024). Initially, we hypothesized that breastfeeding appeared as a predictor due to its interaction with variables like maternal age and Cesarean delivery, both linked to ASD. However, findings from the current research reveal that breastfeeding is not merely a proxy for these factors but an independent variable. Furthermore, this study advances knowledge by examining breastfeeding in a nuanced manner—beyond a binary yes/no framework—to include duration and age-limited feeding patterns. These variables underscore the higher prevalence of exclusive formula use and the earlier cessation of breastfeeding among children with ASD.

Breastfeeding is a shared mother-infant activity influenced by both maternal and infant factors. Our study found that breastfeeding durations are generally shorter in infants with ASD compared to TD infants. These findings do not suggest a causal relationship but rather reflect early ASD manifestations, such as reduced social interactions and emotion dysregulation (Brian et al., 2015; Lemcke et al., 2018; Soke et al., 2019; Varma & de Souza, 2023; Zwaigenbaum et al., 2013). Given that many mothers face significant difficulties in breastfeeding despite their intention to continue, we advocate for healthcare providers, including lactation consultants, to approach these challenges with an understanding of potential ASD-related factors. Additionally, we identified an interaction between SES and breastfeeding, particularly during the 6–12 month period, as well as by exclusive breastfeeding and formula-feeding practices. This nuanced relationship warrants further investigation to better understand its underlying drivers and implications.

## Supporting information

Supplemental Breastfeeding Data Figures

## Data Availability

All data in the present study belong to third-party government agencies and require direct request for approval.

## Acknowledgments

Thank you to Mr. Roe Cohen and Dr. Galit Shefer from TIMNA-Israel Ministry of Health’s Big Data Platform, Ministry of Health, Jerusalem, Israel for their assistance with database management. We wish to thank Dr. Dina Zimmerman, MoH, for her input regarding the interpretation of baby-wellness records. We appreciate Liat Nativ for the assistance with data management.

## Footnotes

1 Top age range is not included. For example, the 0-3 includes data from birth to 2 months and 30 days.

