## Supplemental Breastfeeding Data Figures for "The Interplay Between Breastfeeding and Autism Spectrum"

**Supplemental Materials**

**Figure S1.** Breastfeeding Duration by child's ethnicity, birth type, maternal age groups

Figure S1a. Breastfeeding Duration by Child's Ethnicity

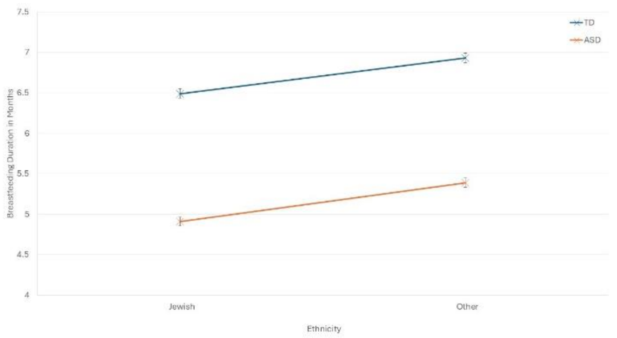


*Note.* Other includes Arab Muslim, Druze, Muslim Bedouin and Arab Christian.

Figure S1b. Breastfeeding Duration by Birth Type

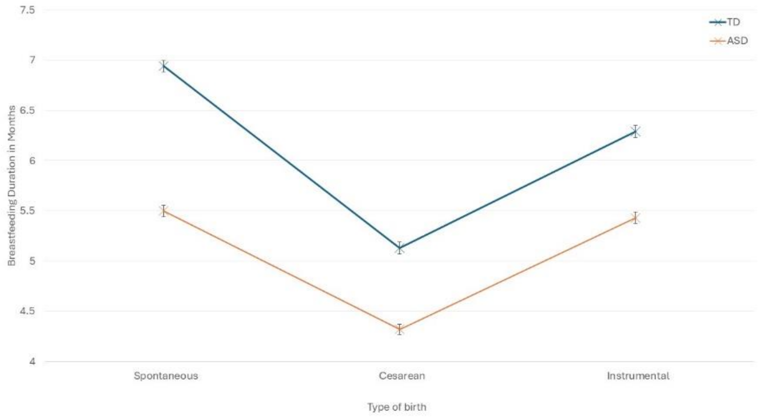

Figure S1c. Breastfeeding duration by Maternal Age Group

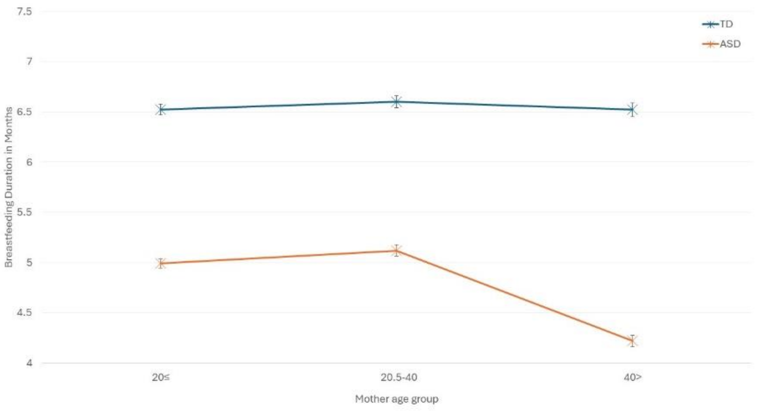


**Figure S2. Percentage of Children in Feeding Type by Group and SES**

Figure S2a. ASD and SES 0-3 months

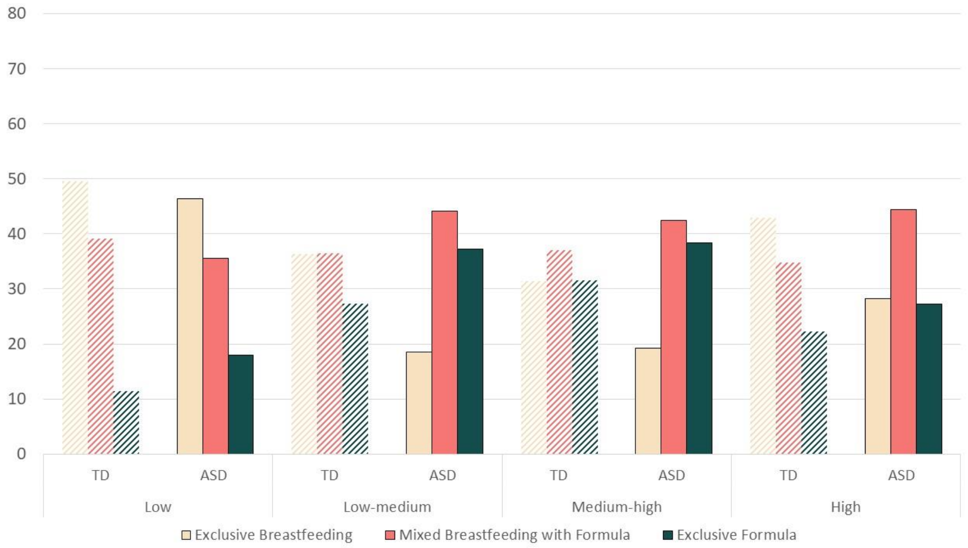


Figure S2b. ASD and SES 3-6 months

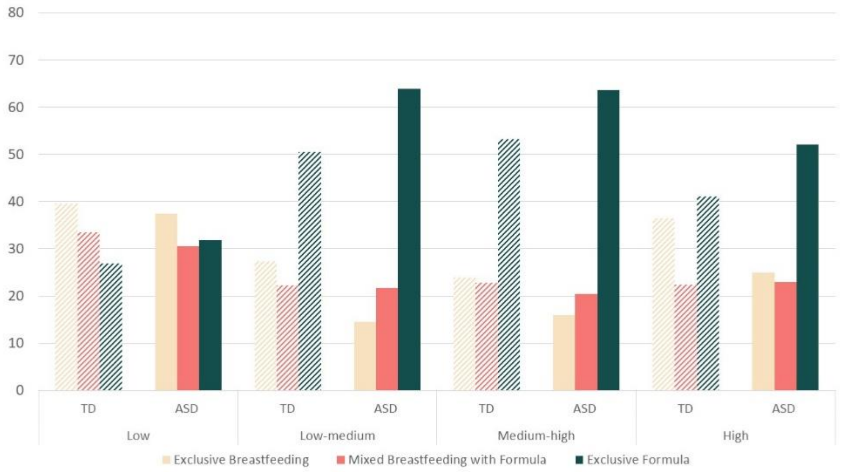

Figure S2c. ASD and SES 6-12 months

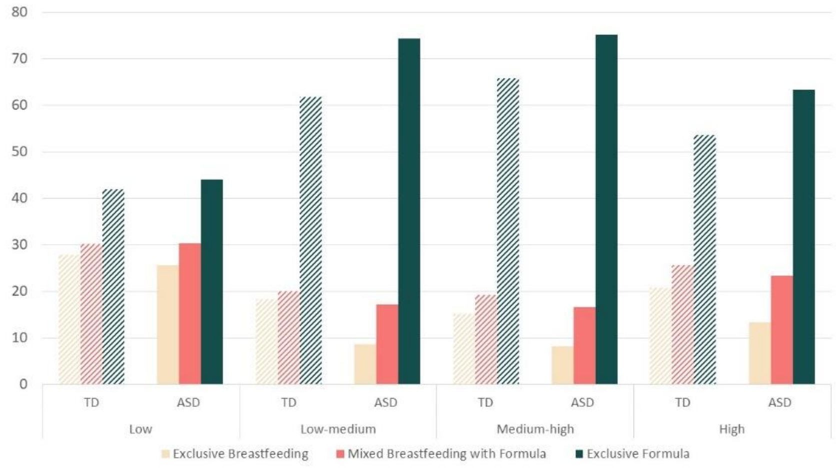
